# Longitudinal Tracking of Valve Regurgitation from Peripheral Photoplethysmography Using a 1D Hemodynamic Surrogate: a Joint MCMC-Kalman Framework

**DOI:** 10.64898/2026.04.30.26352164

**Authors:** Kiran Bhattacharyya

**Affiliations:** SKA Labs Atlanta, GA, USA

**Keywords:** photoplethysmography, valvular heart disease, aortic regurgitation, mitral regurgitation, 1D hemodynamics, Bayesian inference, Kalman filtering, inverse problem

## Abstract

Aortic (AR) and mitral (MR) regurgitation are progressive valvulopathies whose management depends on tracking severity over months to years, currently via infrequent clinic-bound echocardiography. We investigated whether peripheral photoplethysmography (PPG) can track AR and MR longitudinally despite unknown day-to-day hemodynamic variation. A one-dimensional Navier–Stokes arterial model from aortic root to pedal digital artery, with parametric AR and MR at the inlet, generated distal waveforms from which we extracted eleven morphological features. Univariate sensitivity, Cramér– Rao bound, and feature-ablation analyses quantified theoretical observability under realistic noise and physiological variability. A neural surrogate of the 1D model was embedded in a joint Markov Chain Monte Carlo (MCMC)–Kalman scheme that tracked regurgitant fraction monthly while marginalizing over unknown systemic parameters. Across 240 synthetic patients spanning stable and high-risk AR and MR, with and without progressive hypertension, over 36 months, the tracker achieved RMSE of 0.023–0.028 for AR and 0.075–0.101 for MR in regurgitant-fraction units. Dicrotic notch timing was most informative for both valves; augmentation index was uniquely critical for MR. Progressive hypertension paradoxically improved MR tracking by 16–18%. MCMC-Kalman tracking errors arose mainly from over-constrained latent parameters. These results support the theoretical feasibility of wearable PPG-based surveillance of valve regurgitation and motivate prospective clinical validation.

## 1 Introduction

Valvular heart disease affects roughly 2.5% of the general adult population and its prevalence rises steeply beyond age 65 [1, 2, 3]. Aortic regurgitation (AR) and mitral regurgitation (MR) together account for a substantial share of this burden: MR is the second most common single-valve lesion in industrialized countries, and AR, although less prevalent, carries comparable long-term morbidity once it becomes clinically significant [2, 3]. Both lesions are typically chronic and slowly progressive with hemodynamic consequences. Changes unfold over months to years before decompensation [4, 5, 6].

Due to this slow trajectory, AR and MR management is fundamentally a *surveillance* problem. Contemporary guidelines recommend periodic re-evaluation of asymptomatic patients, typically every 6 to 24 months, depending on severity. Clinical intervention generally occurs at onset of symptoms, the crossing of specific regurgitant-fraction or orifice-area thresholds, or evidence of progressive ventricular remodeling [5, 6]. Data indicate that the rate of progression varies substantially across individuals and can accelerate abruptly (for example, with chordal rupture or cusp prolapse), so that the interval between surveillance visits often exceeds the clinically relevant timescale of deterioration [4]. The longitudinal trajectory of the regurgitant fraction is what ultimately determines the timing of intervention.

The current clinical standard for this surveillance is transthoracic echocardiography (TTE) but transesophageal echocardiography or cardiovascular magnetic resonance are used as well [7, 5]. Although TTE provides high-fidelity quantification of regurgitant volume, effective orifice area, and chamber remodeling, it is intrinsically episodic: each measurement requires a trained sonographer, specialized equipment, and a clinic visit, and successive measurements are separated by months during which disease progression is unobserved. Inter- and intra-observer variability in regurgitation severity grading further complicates the detection of slow drifts over time [7]. There is therefore a strong clinical motivation for complementary, low-burden modalities that can provide a denser, continuous estimate of valvular status between echocardiographic visits.

Photoplethysmography (PPG) is an appealing candidate for such a modality. Optical PPG sensors are already embedded in smartphones, smartwatches, rings, and clinical pulse oximeters, providing passive and continuous recordings of the peripheral blood volume pulse at minimal cost [8, 9, 10]. The morphology of the peripheral PPG waveform encodes a rich combination of cardiac output, arterial compliance, wave reflection, and ventricular–arterial coupling, and has been exploited extensively to estimate heart rate, respiratory rate, oxygen saturation, arterial stiffness, vascular age, and even blood pressure with some success [11, 12, 13, 14, 15, 16, 17]. PPG has also been shown, in numerical and clinical studies, to respond sensitively to systemic hemodynamic perturbations such as mental stress and arterial aging [18, 19]. Whether and how it can be used to track the progression of a specific valvular lesion, however, remains essentially unexplored: recent AI-ECG pipelines have demonstrated that surface electrocardiograms carry detectable signatures of moderate-to-severe AR and MR [20], but analogous tools for the peripheral pulse wave are lacking.

Bridging wearable PPG signals to a specific pathophysiological quantity such as regurgitant fraction requires an explicit biophysical link between valvular dynamics and distal waveform morphology. One-dimensional (1D) models of pulse-wave propagation, based on the Navier–Stokes equations in compliant tubes with Windkessel terminations, provide such a link: they have been extensively validated against in vivo and in vitro measurements and reproduce the key features of peripheral pressure, flow, and volume waveforms under healthy and pathological conditions [21, 22, 23, 24, 25, 26, 27, 18]. Inverting such models to recover unobserved physiological parameters from peripheral measurements is a classical inverse problem, tackled previously with variational, sequential, and Bayesian data-assimilation approaches [28, 29, 30, 31, 32, 33]. A longstanding challenge in this setting is that the unknown circulatory parameters are strongly coupled and only partially identifiable from a single waveform snapshot; longitudinal observation and informative priors are typically required to disambiguate them.

In this work, we address the specific question of whether the longitudinal progression of AR and MR fractions can be recovered from peripheral PPG morphology alone, despite unknown and drifting background hemodynamics. Our contribution is threefold. First, we use a 1D Navier–Stokes arterial model with explicit parametric representations of AR and MR at the aortic inlet to generate physiologically realistic peripheral waveforms, from which eleven morphological PPG features are extracted and characterized. Second, we quantify the theoretical observability of each regurgitation fraction through univariate sensitivity, Cramér–Rao Bound, and feature-ablation analyses, establishing which PPG features carry valve-specific information and how many independent measurements are required to exceed a clinically relevant precision threshold under realistic physiological variability. Third, we embed a trained neural surrogate of the 1D model within a joint MCMC–Kalman inference framework that tracks the regurgitant fraction month-by-month while jointly marginalizing over the latent systemic circulation parameters, warm-starting the MCMC ensemble from the previous month’s posterior and fusing its output with a Kalman predictor of disease progression. We demonstrate the approach in two validation studies: a small set of canonical single-patient trajectories that exercise stable, accelerating, and decompensating disease dynamics, and a population-scale study of 240 synthetic patients across eight clinical scenarios, including progressive hypertension confounders designed to stress-test the tracker’s ability to separate valvular decline from concurrent systemic remodeling. Together, these analyses establish the theoretical ceiling and practical limits of PPG-based longitudinal regurgitation tracking and provide a concrete computational scaffold for translation into wearable surveillance of valvular heart disease.

## 2 Methods

### 2.1 Hemodynamic Model

We represented the arterial pathway from the aortic root to the pedal digital artery as a single one-dimensional (1D) compliant tube. The resulting hemodynamics are governed by the nonlinear 1D Navier–Stokes equations, solved with a Lax–Friedrichs scheme, closed by an elastic tube law for wall mechanics and a three-element Windkessel boundary condition for the terminal microcirculation. A single-pathway 1D formulation is a well-established reduced-order approximation in pulse-wave hemodynamics and is known to reproduce the principal features of peripheral pressure, flow, and volume waveforms observed both in vitro and in vivo [21, 22, 24, 23, 26, 25, 27, 18, 34]. It is therefore well-suited to establishing feasibility bounds for linking peripheral PPG morphology to valvular biophysical parameters.

#### Governing Equations

The model solves the 1D Navier-Stokes equations for blood flow in compliant vessels, consisting of conservation of mass and momentum:

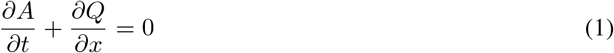

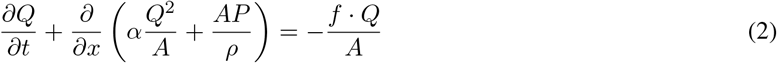

where *A*(*x, t*) is the vessel cross-sectional area [m^2^], *Q*(*x, t*) is the volumetric flow rate [m^3^*/*s], *P* (*x, t*) is the transmural pressure [Pa], *ρ* = 1060 kg*/*m^3^ is blood density, *α* = 1.0 is the momentum correction coefficient (flat velocity profile), and *f* represents viscous friction forces.

#### Constitutive Relations

Vessel wall mechanics are described by a nonlinear tube law relating pressure to area:

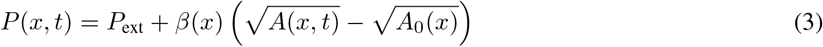

where *P*_ext_ = 0 Pa is external pressure, *A*_0_(*x*) is the reference cross-sectional area at zero transmural pressure, and *β*(*x*) [Pa *·* m] is the wall stiffness parameter. The local pulse wave velocity is:

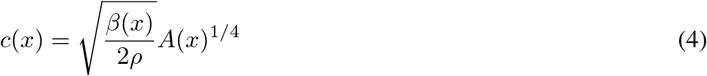

Viscous friction follows a Poiseuille-like profile:

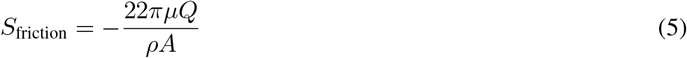

where *µ* = 0.004 Pa *·* s is blood dynamic viscosity.

#### Arterial Geometry

The computational domain spans *L* = 1.0 m from the proximal aorta to the pedal digital artery, discretized with spatial step ∆*x* = 0.01 m. The baseline geometry features exponential tapering from proximal radius 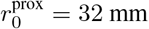 to distal radius 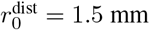:

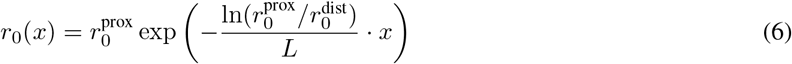

Wall stiffness increases distally to reflect physiological arterial stiffening, ranging from *β*^prox^ = 5 *×* 10^4^ Pa *·* m (yielding PWV *≈* 5 m/s in the aorta) to *β*^dist^ = 2 *×* 10^6^ Pa *·* m (PWV *≈* 15 m/s in pedal digital arteries).

#### Numerical Method

The system is solved using the Lax-Friedrichs finite difference scheme, chosen for its stability properties. The time step satisfies the Courant-Friedrichs-Lewy (CFL) condition with CFL number 0.4:

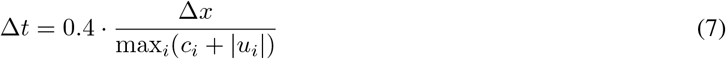

At the inlet (proximal aorta), a physiologically realistic cardiac flow waveform is prescribed when simulating normative valve function, consisting of rapid systolic ejection (35% of cycle), brief early diastolic backflow representing valve closure, and zero diastolic flow. Stroke volume (baseline 70 mL) is adjusted based on mean arterial pressure (MAP) with sensitivity of −1% per mmHg deviation from 90 mmHg baseline. Pathological valve regurgitation is modeled by altering this inlet waveform as described below in Section 2.2.

At the outlet (pedal digital artery), a three-element Windkessel model (*R*_1_–*C*–*R*_2_) represents peripheral impedance, with characteristic impedance *R*_1_ = 5 *×*10^7^ Pa *·* s*/*m^3^, peripheral resistance *R*_2_ = 5 *×*10^8^ Pa *·* s*/*m^3^, and compliance *C* = 2 *×* 10^*−*9^ m^3^*/*Pa. The resulting distal cross-sectional area waveform reproduces the canonical features of an optically-acquired PPG pulse — a rapid systolic upstroke, a dicrotic notch, and an exponential diastolic decay [8, 9, 10] — across a physiological range of mean arterial pressures (Appendix **??**).

### 2.2 Mathematical Representation of Valve Regurgitation in the Hemodynamic Model

Valve regurgitation is represented by modifying the proximal aortic inlet waveform, *Q*_in_(*t*), together with physiologically consistent adjustments to the systemic afterload [34, 5, 6]. Under baseline conditions, *Q*_in_(*t*) consists of a rapid systolic sinusoidal-style ejection phase, a brief early-diastolic backflow corresponding to aortic valve closure, and zero subsequent diastolic flow. The baseline forward stroke volume (SV_base_) scales inversely with mean arterial pressure (MAP) to capture physiological afterload sensitivity. Below, we describe how AR and MR are each imposed on this baseline waveform; the specific mathematical forms are motivated by the hemodynamic phenotypes described in standard echocardiographic and clinical references [7, 4, 34].

#### Mitral Regurgitation (MR)

Mitral regurgitation is modeled exclusively as a modification to the forward systolic ejection profile. Because a fraction of ventricular stroke volume is ejected backward into the lower-impedance left atrium rather than into the aorta, the effective forward stroke volume is reduced [4, 7]. The gross forward volume is scaled by a non-linear compensation factor that depends on the MR fraction (*f*_MR_):

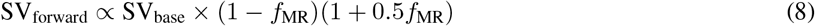

Because the low-resistance atrial pathway accelerates ventricular emptying, the temporal landmarks of systole also shift [34]. The time to peak systolic flow (*t*_peak_) is shifted earlier within the cardiac cycle (duration *T*_cycle_) by a fractional MR onset delay (*d*_MR_), while the absolute duration of the systolic ejection period (*t*_end_sys_) is truncated in proportion to regurgitation severity:

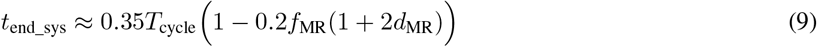

The net effect is a steeper, narrower systolic ejection with a sharp initial wavefront and a lower peak forward pressure.

#### Aortic Regurgitation (AR)

Unlike MR, aortic regurgitation disrupts both systolic and diastolic phases of the aortic flow waveform. In chronic, compensated AR, eccentric hypertrophy augments the gross forward stroke volume so that net cardiac output is preserved despite ongoing diastolic losses [5, 6, 34]. We capture this systolic compensation by scaling the forward ejected volume through an asymptotic bounded function of the AR fraction (*f*_AR_):

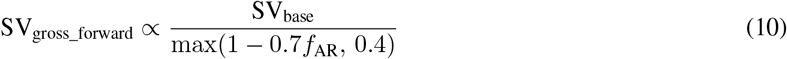

During diastole (*t > t*_end_sys_), the incompetent aortic valve permits continuous regurgitant backflow from the aorta into the left ventricle. We model this as an exponentially decaying negative flow profile extending throughout the diastolic interval:

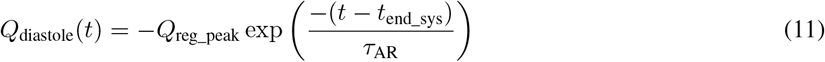

where *τ*_AR_ is a patient-specific decay constant encoding the regurgitant orifice geometry and pressure-gradient collapse. The peak regurgitant flow *Q*_reg_peak_ is constrained analytically so that the integral of the exponential decay over diastole matches the target regurgitant loss volume (*V*_reg_ = *f*_AR_ *×* SV_gross_forward_).

Severe AR is clinically associated with peripheral vasodilation, an autoregulatory response that favors forward flow over regurgitation down the aortic–ventricular gradient [5, 34]. We enforce this in the 1D model by reducing the terminal Windkessel resistance *R*_2_ as a function of AR severity:

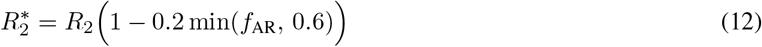

Together with the augmented forward stroke volume, this systemic impedance reduction produces the classical “water-hammer” widened pulse pressure characteristic of severe AR.

### 2.3 Algorithmic Extraction of Morphological PPG Features

We extracted morphological features from the terminal cardiac cycle of the simulated distal arterial cross-sectional area waveform (*A*_dist_(*t*)), which serves as a biomechanical proxy for the optical PPG volume signal [8, 10]. The corresponding proximal driving pressure (*P*_prox_(*t*)) was evaluated concurrently to extract propagation metrics. Before feature localization, both waveforms were linearly detrended to remove baseline wander and normalized to zero mean and unit variance, preserving relative morphology while standardizing amplitudes.

The feature set of eleven metrics was chosen to span the two established families of PPG contour analysis: generalized markers of cardiovascular stiffness and wave reflection [11, 9, 16, 15, 17, 19], and targeted signatures of valvular regurgitation.

#### Propagation and Generalized Stiffness Features

- **Pulse Transit Time (PTT) & Pulse Wave Velocity (PWV):** PTT was calculated by foot-to-foot cross-correlation, with the fiducial foot identified as the local signal minimum immediately preceding the maximum gradient of the systolic upstroke [12]. The effective pulse-wave velocity (PWV_eff_) was obtained by dividing the fixed 1D domain length (*L*) by the PTT.
- **Augmentation Index (AI):** To avoid the noise sensitivity of second-derivative inflection points [11, 9], AI was estimated via a robust proxy that tracks post-peak systolic decay. The waveform was sampled at 20% and 40% of the diastolic interval; slower decay indicates greater influence from backward-traveling reflections and therefore a higher AI.
- **Reflection Index (RI):** An area-based metric for distal wave reflection, obtained by partitioning the waveform at the systolic peak (*t*_sys_). RI is the ratio of the diastolic-phase integral to the total area bounded under the baseline-shifted cycle.
- **Systolic Time Ratio & Dicrotic Notch Timing:** The systolic time ratio is the fraction of the cardiac cycle required to reach the maximum peak (*t*_sys_*/T*_cycle_). Dicrotic notch timing is the elapsed time from the systolic peak to the first prominent post-systolic negative inflection, identified via gradient zero-crossings and corresponding to aortic valve closure [9].

#### Regurgitation-Targeted Features

Reflecting the distinct hemodynamic perturbations imposed by incompetent valves [7, 34], four additional features were defined to isolate diastolic runoff and pulse-pressure widening:

- **Diastolic Decay Rate:** Quantifies the pathological “runoff” of systemic pressure characteristic of severe AR. The late-diastolic tail was isolated and an exponential decay constant was estimated by ordinary least-squares regression of the log-transformed signal against time.
- **Pulse Pressure Index:** Captures the “water-hammer” widened pulse pressure of compensated AR. Defined as the peak-to-trough systemic pulse pressure normalized by the margin of mean pressure above the diastolic baseline, (*P*_sys_ *− P*_dia_) */* (*P*_mean_ *− P*_dia_).
- **Dicrotic Notch Depth:** Because worsening AR blunts or obliterates the aortic-closure notch, we track the amplitude of the dicrotic minimum as a fraction of the bounding systolic peak and local diastolic floor.
- **Diastolic Area Fraction:** Conceptually related to RI, but computed on the absolute, un-normalized cross-sectional area curves. Incompetent mitral or aortic valves redistribute intravascular volume away from the distal bed during diastole, reducing this ratio.

#### 2.3.1 Measurement Noise and Uncertainty Configuration for PPG Features

To generate realistic synthetic observations, each PPG feature was corrupted by additive Gaussian noise 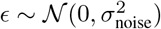. The noise level *σ*_noise_ lumps together three sources of variability commonly observed in wearable PPG acquisition: optical instrument variability, motion artifact, and algorithmic ambiguity during fiducial-point extraction [8, 10]. Baseline standard deviations were assigned per feature based on clinical ranges and mathematical robustness:

- **Temporal Landmarks:** Pulse Transit Time (PTT, *σ* = 2.0 ms) and Dicrotic Notch Timing (*σ* = 5.0 ms). These uncertainties reflect baseline jitter equivalent to a standard 200–500 Hz acquisition sampling rate limit compounded by smoothing-induced latency applied during gradient-based localized foot/notch inflection identification.
- **Area and Ratio Morphometrics:** Reflection Index (RI, *σ* = 0.03), Augmentation Index (AI, *σ* = 0.03), Systolic Time Ratio (*σ* = 0.015), and Diastolic Area Fraction (*σ* = 0.03). Because these are intrinsically self-normalized, geometrically bounded, unit-less fractions, they exhibit resilience against baseline wandering and absolute amplitude miscalibration. Consequently, they are assigned tight variability margins.
- **Curve-Fitting Parameters:** Diastolic Decay Rate (*σ* = 0.35 s^*−*1^). This metric relies upon logarithmic linear regression executed strictly over the low-amplitude late-diastolic tail. Because optical signal-to-noise ratio (SNR) is at its poorest geometric minimum during this phase, this value is subject to broader variance.
- **Magnitude Indices:** Pulse Pressure Index (*σ* = 0.07) and Dicrotic Notch Depth (*σ* = 0.04). These parameters evaluate vertical pulse extremes, and their variance reflects the fundamental limits of transducer linearity and minor venous pooling oscillations.

### 2.4 Parameters defining systemic circulation

The 8 parameters that characterize systemic circulation 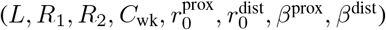 are referred to as *θ*_circ_ and their values as used in our work are provided in the columns of Table 1. We report which values are used and why for each experiment.

**Table 1:** Ranges for Systemic Circulation Parameters (*θ*_circ_) and Usage Across Experiments.

| Parameter | Units | Bounds | Nominal<sup>†</sup> | Nominal ± noise<sup>‡</sup> |
| --- | --- | --- | --- | --- |
| $L$ (Length) | m | [0.7, 1.3] | 0.95 | $\pm 10\%$ |
| $R_1$ (Prox. Resistance) | Pa·s/m<sup>3</sup> | $[2 \times 10^7, 1 \times 10^8]$ | $4.47 \times 10^7$ | $\pm 20\%$ |
| $R_2$ (Dist. Resistance) | Pa·s/m<sup>3</sup> | $[1 \times 10^8, 1 \times 10^9]$ | $3.16 \times 10^8$ | $\pm 40\%$ |
| $C_{wk}$ (Windkessel Comp.) | m<sup>3</sup>/Pa | $[5 \times 10^{-10}, 5 \times 10^{-9}]$ | $1.58 \times 10^{-9}$ | $\pm 20\%$ |
| $r_{0,p}$ (Prox. Radius) | m | [0.008, 0.016] | 0.0113 | $\pm 10\%$ |
| $r_{0,d}$ (Dist. Radius) | m | [0.001, 0.002] | 0.0014 | $\pm 10\%$ |
| $\beta_p$ (Prox. Stiffness) | Pa | $[2 \times 10^4, 1 \times 10^5]$ | $4.47 \times 10^4$ | $\pm 25\%$ |
| $\beta_d$ (Dist. Stiffness) | Pa | $[8 \times 10^5, 4 \times 10^6]$ | $1.79 \times 10^6$ | $\pm 25\%$ |

<sup>†</sup> Nominal values are calculated as the geometric mean of the trained physiological bounds: $\theta_{\text{nom}} = \sqrt{\theta_{\text{min}} \cdot \theta_{\text{max}}}$ and used for a representative individual.
<sup>‡</sup> We use these ranges to simulate the biological variance of $\theta_{\text{circ}}$ within a representative individual.

### 2.5 Sensitivity Analysis of Peripheral Pulse Features to Valve Regurgitation

To assess the theoretical detectability of AR and MR from peripheral PPG features under idealized conditions, we performed a univariate sensitivity analysis. The hemodynamic solver was evaluated on a grid of 25 linearly spaced regurgitation fractions (0 *≤ θ ≤* 0.5) for AR and MR independently. During each sweep, the complementary regurgitant fraction was held at zero, and the remaining hemodynamics were fixed at the nominal *θ*_circ_ values (Table 1), a heart rate of 70 bpm, and a mean arterial pressure of 95 mmHg.

For each of the 11 morphological features *f*_*i*_ (Section 2.3), the linear sensitivity to the regurgitant fraction was quantified via ordinary least-squares regression across the sweep, yielding the partial derivatives *∂f*_*i*_*/∂*AR and *∂f*_*i*_*/∂*MR. To account for heterogeneous units and feature scales, sensitivities were normalized by the modality-specific noise standard deviation *σ*_noise,*i*_ (Section 2.3.1). The resulting signal-to-noise ratio, |*∂f*_*i*_*/∂θ*| */σ*_noise,*i*_, serves as a feature-level proxy for the theoretical observability of regurgitation severity when all other hemodynamic parameters are held constant.

### 2.6 Theoretical Precision Limits Under Physiological Variation

Clinical translation requires robustness to day-to-day physiological variability. We therefore evaluated theoretical lower bounds on the estimation variance of the AR and MR fractions using the Cramér–Rao Bound (CRB) framework [33]. The Fisher Information Matrix (FIM) was computed for *N* observations in which longitudinal variability was simulated by sampling *θ*_circ_ uniformly from Nominal*±* noise, heart rate from *𝒰* (60, 90) bpm, and MAP from *𝒰* (80, 110) mmHg.

For each perturbed state, the local Jacobians mapping the regurgitation parameters to the 11 PPG features were retrieved from the hemodynamic model. The FIM was aggregated over the *N* observations and weighted by the inverse feature-noise covariance matrix. Two complementary analyses were performed: (i) the CRB standard deviations *σ*_AR_ and *σ*_MR_ were evaluated at three sample sizes (*N ∈ {*10, 25, 50*}*) while the baseline measurement noise was scaled by multipliers *m ∈ {*0.5, 1.0, 2.0, 4.0 *}*; and (ii) *N* was continuously swept from 2 to 100 at baseline noise, in order to determine the longitudinal data requirements needed to drive tracking uncertainty below a 5% margin.

### 2.7 Feature Ablation and Importance Profiling

To quantify the contribution of individual features to overall estimation precision, we performed a feature-ablation study within the same CRB framework. Using *N* = 50 physiological samples generated as in Section 2.6, we first computed an unablated baseline FIM yielding reference standard deviations *σ*_AR_ and *σ*_MR_.

Each feature was then ablated in turn by inflating its measurement noise variance by a factor of 10, and the CRB was re-evaluated. This simulates the effect of rendering that feature effectively unusable through motion artifact, poor signal quality, or algorithmic failure [10]. Features were ranked by the composite loss in precision, defined as the average increase in *σ*_AR_ and *σ*_MR_ relative to baseline, isolating the subset most essential for recovering valvular signatures under physiological variability.

### 2.8 Surrogate Neural Network Training and Architecture

Sequential Bayesian tracking requires thousands of forward-model evaluations per monthly update, making direct use of the 1D solver computationally impractical. We therefore trained a feed-forward neural network as a differentiable surrogate of the 1D hemodynamic forward map, a standard strategy for accelerating inverse problems in computational hemodynamics and digital-twin applications [31, 32, 35].

#### 2.8.1 Synthetic Data Generation

The surrogate approximates a 14-dimensional to 11-dimensional map. The input vector Θ *∈* ℝ^14^ consists of four valvular regulators (AR fraction, MR fraction, AR diastolic decay constant *τ*, and MR systolic onset delay) and ten generalized cardiovascular parameters (heart rate, MAP, and the eight *θ*_circ_ parameters).

To cover the joint parameter space uniformly, training samples were drawn via Latin Hypercube Sampling (LHS) over population-wide bounds: AR and MR fractions from 0.0 (competent) to 0.6 (severe), heart rate 45–150 bpm, MAP 50–200 mmHg, and *θ*_circ_ within the Bounds listed in Table 1. For each draw, the 1D solver produced the corresponding 11-dimensional output *f* (Θ) combining stiffness, reflection, and regurgitation-specific features.

#### 2.8.2 Network Architecture and Pre-processing

Because the 14 input parameters and 11 output metrics span widely different scales, both the input matrix **X** and target matrix **y** were z-score normalized to zero mean and unit variance using the global statistics of the training set.

The surrogate is a multi-layer perceptron with an input layer, three fully connected hidden layers of 128, 128, and 64 neurons, and an 11-neuron linear readout. Each hidden layer uses a Rectified Linear Unit (ReLU) activation followed by dropout (*p* = 0.1) for regularization.

#### 2.8.3 Training Regimen and Early Stopping

The network was trained by minimizing the mean squared error (MSE) with the Adam optimizer (initial learning rate 10^*−*3^) and mini-batches of 64. The training set was split 90%/10% into training and validation subsets, and training ran for up to 400 epochs with early stopping after 40 epochs without validation-loss improvement.

Generalization was assessed on 5,000 held-out samples drawn under the same LHS protocol (Section 2.8.1); we report the RMSE and coefficient of determination (*R*^2^) for each of the 11 PPG features.

#### 2.8.4 Use of the Neural Surrogate

The trained surrogate is used exclusively to solve the inverse problem during longitudinal tracking. At each monthly time step, synthetic PPG observations are generated by the full 1D solver using the (unknown to the tracker) ground-truth patient parameters; the surrogate then replaces the solver *only* during Bayesian inference of the latent hemodynamic inputs, as described in Sections 2.10 and 2.11. This separation prevents inference and generation from sharing the same approximation error.

### 2.9 Longitudinal Bayesian Estimation via MCMC-Kalman Filtering

To track the progression of valve regurgitation severity against unknown drifts in *θ*_circ_, we implemented a hybrid Markov Chain Monte Carlo (MCMC) and Kalman filtering framework. Similar hybrid data-assimilation strategies have been successfully deployed in computational hemodynamics to separate slowly-varying patient parameters from faster-varying pathological states [29, 31, 28, 32]. In our implementation, the tracker maintains a joint posterior over the regurgitant fraction (either AR or MR) and the eight *θ*_circ_ parameters at each time step.

#### 2.9.1 Joint-log Posterior and Kalman Framework

At each time step *t*, the patient generates *N* noisy PPG observations **f**_*t*_ =*{f*_*t*,1_, …, *f*_*t,N*_*}*. Each observation is drawn under random sampling of heart rate, MAP, and *θ*_circ_ from the intra-individual variance range (Nominal*±* noise, Table 1), together with beat-to-beat jitter (*±*0.03) in the AR/MR fraction *V*_*t*_. AR and MR scenarios were simulated separately rather than concurrently. The joint log-posterior over the unknown parameters *V*_*t*_ and *θ*_circ_ is:

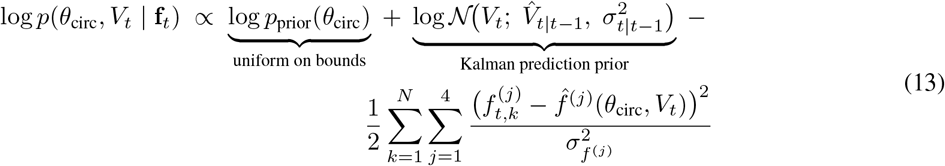

where 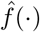(*·*) denotes the surrogate prediction and 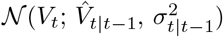 is the one-step-ahead Kalman prediction prior.

The temporal evolution of *V*_*t*_ is modeled as a Kalman-style predict–update cycle. The predictive prior at time *t* is obtained by propagating the posterior at *t −* ∆*t* forward:

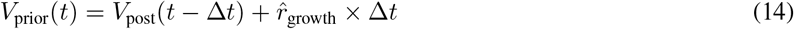

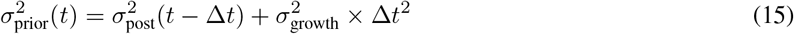

where 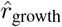 is the adaptively estimated progression rate and *σ*_growth_ = 0.02 /month represents the process noise of the progression model. The posterior is then obtained by Bayesian fusion of the predictive prior with the current MCMC estimate:

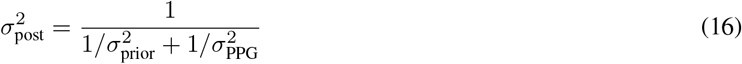

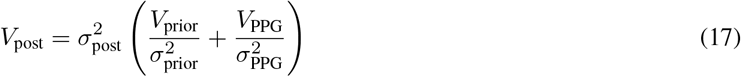

where *V*_PPG_ and 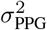 are the MCMC-derived posterior mean and variance of *V*_*t*_ from the current month’s observations (Section 2.9.4). The progression rate is updated online via an exponential moving average with learning rate *α* = 0.3:

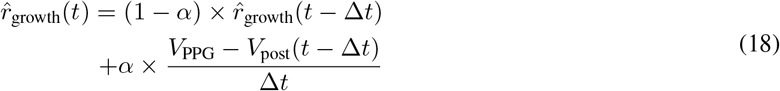

Crucially, the likelihood in Equation 13 sums over the *current month’s measurements only*. Information about *θ*_circ_ from earlier months is carried forward implicitly through the MCMC walker positions rather than through a fixed point estimate, as described next.

#### 2.9.2 Sequential MCMC with Warm-Starting

We sampled the joint posterior (Eq. 13) using the affine-invariant ensemble sampler emcee [36] with 128 walkers. At each subsequent time step (*t >* 1), the ensemble was *warm-started* from the previous month’s final walker positions. A small jitter was applied to relax the ensemble into the updated typical set: 0.5% multiplicative Gaussian noise on *θ*_circ_, and additive noise of scale *σ*_growth_ ∆*t* on *V*_*t*_ to account for expected progression. This yields the sequential Bayesian update:

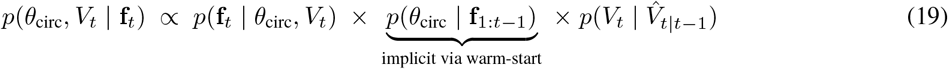

Because the walkers already reside near the previous posterior, no explicit parametric approximation of *p*(*θ*_circ_ |**f**_1:*t* − 1_) is required. Parameter uncertainty narrows or expands organically as longitudinal data accumulate, avoiding the bias that would arise from fixing *θ*_circ_ at a point estimate [32, 33].

#### 2.9.3 Initialization via Log-Space Optimization

At *t* = 1, the walkers must be initialized without access to prior samples. Uniform sampling of the joint space is inefficient because the parameter magnitudes span 17 orders of magnitude (*C*_wk_ *∼* 10^*−*9^ vs. *R*_2_ *∼* 10^8^). We therefore performed 128 multi-start L-BFGS-B optimizations of the negative log-posterior with *θ*_circ_ parameterized in log-space and *V* in natural units. The walkers were then distributed around the top modes with small Gaussian perturbations, providing a well-concentrated initialization near the posterior’s typical set. The first month of MCMC ran for 500 steps; subsequent months used 250 steps with a 150-step burn-in.

#### 2.9.4 Kalman Fusion and Progression Estimation

After discarding burn-in samples, the marginal posterior mean *µ*_ppg_ and standard deviation *σ*_ppg_ of *V*_*t*_ were extracted from the MCMC chain. These serve as the observation (*V*_PPG_, *σ*_PPG_) in the Kalman fusion step (Section 2.9.1), which executes the predict, fuse, and growth-rate updates of Equations 14–18. The fused posterior then seeds the Kalman prediction prior for the next month’s MCMC run, closing the sequential loop.

### 2.10 Simulated Longitudinal Tracking of Regurgitation Severity

We evaluated the tracking framework (Section 2.9) over a 36-month monitoring horizon on canonical disease trajectories representative of chronic and decompensating valve disease [4, 5, 6]. Two overarching scenario classes were examined: constant steady-state progression and accelerated progression. The constant-progression cohort modeled gradual remodeling:

- **Stable AR**: Initiated at an AR fraction of 0.15 with a constant regurgitant fraction progression velocity of *v* = 0.0015*/*month.
- **Stable Primary MR**: Initiated at an MR fraction of 0.18 with an expected progression velocity of *v* = 0.0025*/*month.
- **Secondary MR**: Initiated at an MR fraction of 0.20, representing load-dependent functional disease yielding near-zero underlying anatomical progression (*v* = 0.0) but higher process noise expectation for growth in the tracking system.
- **High-Risk AR**: Modeled aggressive structural decline beginning at a more severe AR fraction of 0.30 (*v* = 0.004*/*month).

The accelerated-progression cohort mirrored the steady-state trajectories until month 6, at which point the progression rate was abruptly increased. Stable AR and Primary MR accelerated to *v* = 0.005/month and *v* = 0.006/month, respectively, representing events such as chordal rupture or flail leaflets [4, 7]. High-Risk AR accelerated to *v* = 0.008/month, and Secondary MR accelerated from *v* = 0.005 *→* 0.008/month with a 2*×* increase in process noise. Critically, the tracker was only given the initial AR/MR fraction and a bounded prior on progression velocity (*σ*_growth_ = 0.02/month, or 0.04/month for Secondary MR); it had no knowledge of the future acceleration.

### 2.11 Population-Scale Validation Across Canonical Disease Scenarios

To characterize tracking accuracy and bias across heterogeneous cohorts, we performed a population-scale validation. The framework was evaluated on 240 independent longitudinal trajectories, evenly divided across eight canonical disease profiles (30 synthetic patients per scenario) over a 36-month horizon:

1. **Stable AR**: Chronic compensated aortic regurgitation with an expected nominal baseline severity of *x*_0_ = 0.15 and a progression velocity of *v*_0_ = 0.0015 month^*−*1^.
2. **Stable MR**: Chronic primary mitral regurgitation characterized by *x*_0_ = 0.18 and *v*_0_ = 0.0025 month^*−*1^.
3. **High-Risk AR**: Aggressive structural decline mapped to a baseline *x*_0_ = 0.30 and escalated velocity *v*_0_ = 0.0040 month^*−*1^.
4. **High-Risk MR**: Evolving flail-leaflet/rapidly remodeling MR mapped to *x*_0_ = 0.30 and velocity *v*_0_ = 0.0060 month^*−*1^.

The remaining four scenarios paired each valvular pathology with a progressive, unobserved hemodynamic confounder (denoted *+HTN*), designed to stress-test the tracker’s ability to separate valvular decline from generalized arterial stiffening:

- **Stable + HTN** (AR and MR): a monotonic +10.0 mmHg rise in MAP over 36 months (+0.28 mmHg/month), together with a 10% relative decrease in peripheral compliance (*C*_*wk*_) and a 10% increase in arterial stiffness (*β*).
- **High-Risk + HTN** (AR and MR): an aggressive +30.0 mmHg rise in MAP (+0.83 mmHg/month), with a 20% reduction in compliance and a 20% increase in stiffness over the same 36 months.

To ensure patient-level statistical independence and reflect population diversity, each synthetic patient’s ground-truth *θ*_circ_ was drawn from the Bounds range (Table 1) and further perturbed within Nominal*±* noise at each monthly observation. The patient’s true initial regurgitation severity was drawn as *x*_0,true_ *∼* 𝒰 (*x*_0_ *−* 0.05, *x*_0_ + 0.05) and the true progression velocity as *v*_0,true_ *∼* 𝒰 (0.7*v*_0_, 1.3*v*_0_). Each synthetic observation additionally jittered the AR/MR fraction by *±* 0.03 to capture beat-to-beat variability. The tracker was initialized at the nominal scenario mean with a bounded prior on progression velocity (*σ*_growth_ = 0.02/month) and had no knowledge of the imposed hypertension or compliance drifts.

Each monthly measurement used 256 synthetic observations. After 36 months of iterative tracking, we computed patient-level root-mean-square error (RMSE) against the randomized ground-truth trajectory and summarized cohort-level fidelity as mean *±* standard deviation within and across scenarios. We additionally examined the relationship between posterior identifiability of the latent *θ*_circ_ parameters and the AR/MR tracking error, to probe the failure modes of joint inference.

## 3 Results

### 3.1 Sensitivity of Peripheral Pulse Features to Regurgitant Fractions

We first evaluated the theoretical sensitivity of the eleven morphological features across a linear sweep of regurgitant fractions (0 *≤ θ ≤* 0.5, Section 2.5), with heart rate, MAP, and *θ*_circ_ held constant to isolate the pathology signature.

The results in Figure 1 reveal a pronounced asymmetry in peripheral observability between AR and MR. AR exerted strong, multi-feature effects on the peripheral waveform. The dominant markers were pulse wave velocity (PWV_eff_) and the tightly coupled stiffness index (slope +6.73 m/s, SNR = 44.86), followed by augmentation index (AI; slope *−*0.71 a.u., SNR = 23.76), pulse transit time (PTT; slope *−*32.93 ms, SNR = 16.46), and dicrotic notch timing (slope *−*50.67 ms, SNR = 10.13).

**Figure 1:**
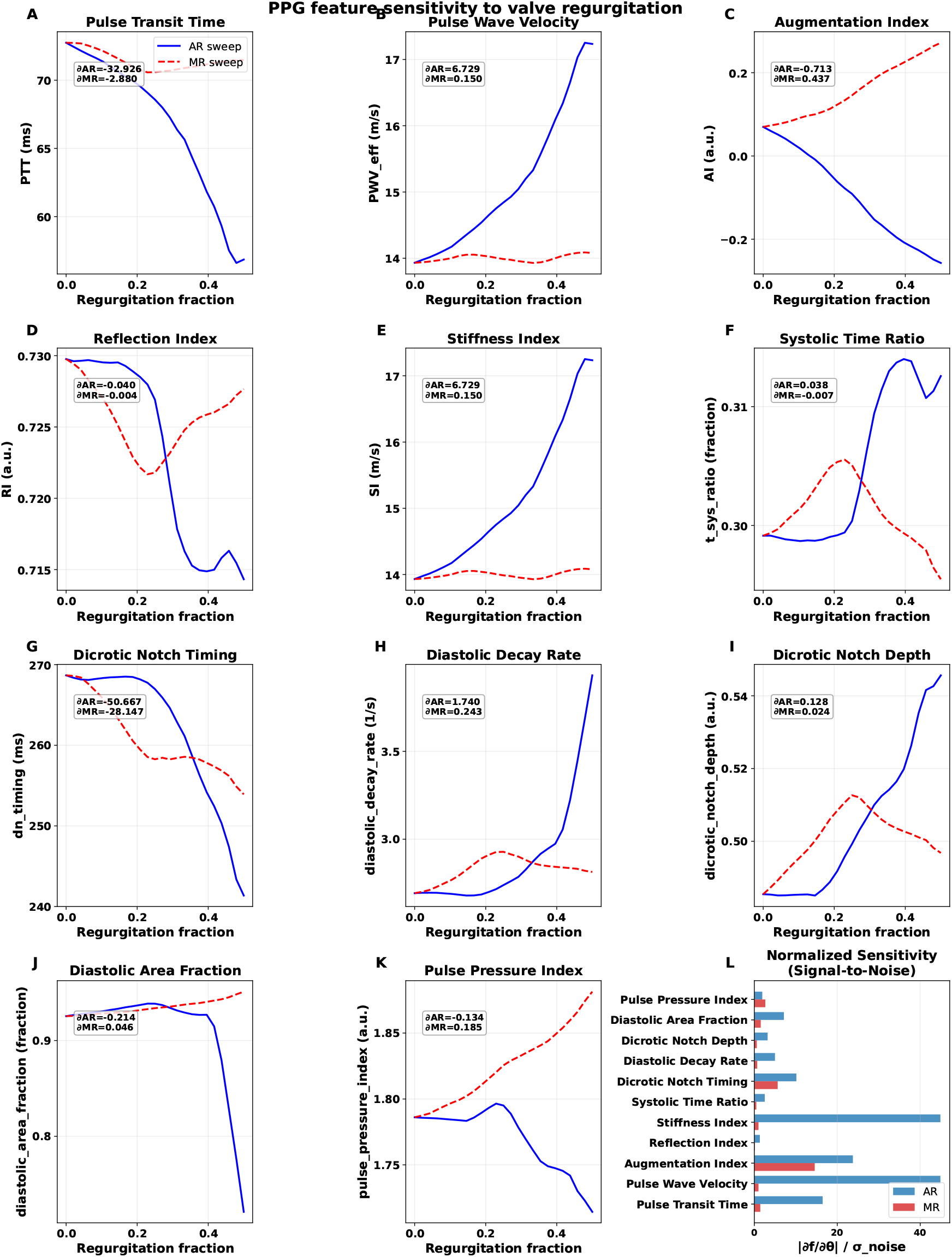
Univariate sensitivity of peripheral photoplethysmography (PPG) waveform features to progressive valve regurgitation. Eleven derived morphological features of the distal pulse wave are evaluated across linearly sweeping aortic regurgitation (AR, solid blue) and mitral regurgitation (MR, dashed red) fractions. Background physiological parameters are held nominally static to isolate the theoretical pathology signature. The aggregate panel (bottom right) presents the normalized Signal-to-Noise Ratio (SNR) for each respective axis (|*∂f/∂θ*| */σ*_noise_), demonstrating high localized observability thresholds for AR in contrast to the heavily muted peripheral footprint of MR.

Pure MR, by contrast, produced a muted peripheral footprint. Only augmentation index exceeded a clinically meaningful SNR (slope +0.44 a.u., SNR = 14.56), with secondary contributions from dicrotic notch timing (SNR = 5.63) and pulse pressure index (SNR = 2.65). Notably, PWV_eff_ and the stiffness index — the dominant AR markers — were essentially insensitive to MR (SNR = 1.00, slope +0.15 m/s).

These univariate SNRs represent an upper bound on observability because they assume a static cardiovascular state. For clinical deployment, the same features must remain informative under intra-individual variation in heart rate, MAP, and *θ*_circ_, which we quantify next.

### 3.2 Theoretical Precision Limits Under Physiological Variation

To probe robustness to day-to-day cardiovascular fluctuations, we computed the Cramér–Rao Bound (CRB) over *N* repeated observations at a nominal disease state (AR = 0.30 or MR = 0.30) with background heart rate, MAP, and *θ*_circ_ drawn from their physiological distributions (Section 2.6).

At baseline PPG feature noise (1.0 *×*, Section 2.3.1), relatively few measurements sufficed to drive the theoretical standard deviation below the 5% clinical margin (Figure 2). With *N* = 10, the CRB was *σ*_AR_ = 0.019 and *σ*_MR_ = 0.037; at *N* = 25, *σ*_AR_ = 0.014 and *σ*_MR_ = 0.017; and at *N* = 50, *σ*_AR_ = 0.009 (0.88%) and *σ*_MR_ = 0.012 (1.19%). The monotonic improvement with *N* reflects the fact that natural physiological variability across measurements averages out non-valvular hemodynamic noise, thereby identifying the valvular component.

**Figure 2:**
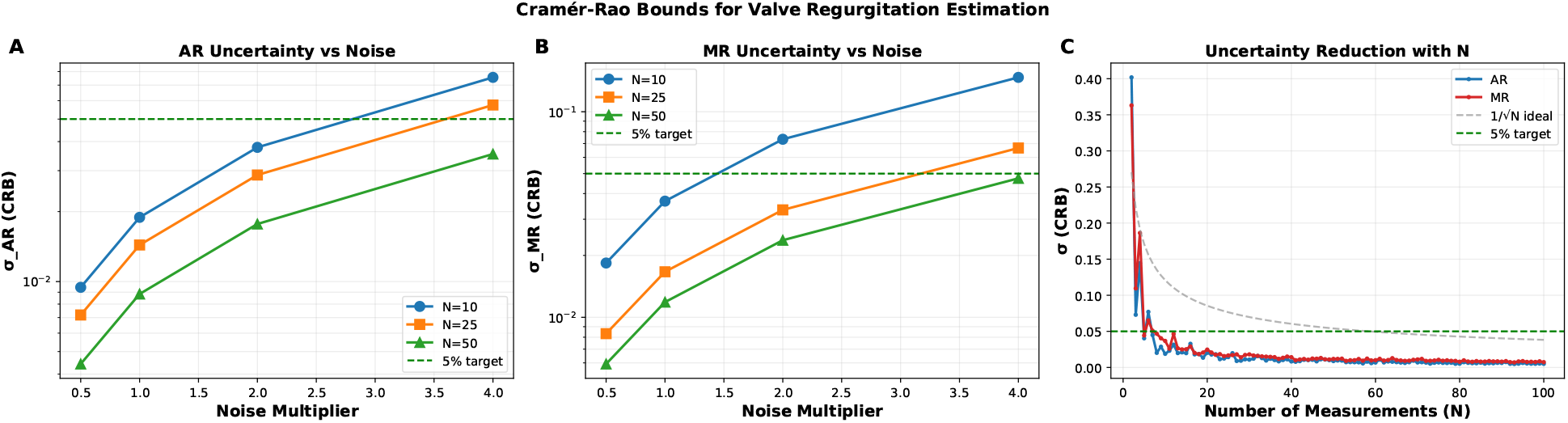
Cramér-Rao Bound (CRB) estimation tracking limits under physiological variance. Theoretical standard deviations for tracking aortic regurgitation (AR) and mitral regurgitation (MR) fractions are computed by aggregating multi-dimensional Fisher Information Matrices. To simulate real-world day-to-day variance, discrete measurements inherently incorporate random physiological perturbations in background heart rate (𝒰 (60, 90) bpm), mean arterial pressure (𝒰 (80, 110) mmHg), and circulatory parameters under Nominal*±* noise conditions (Table 1) **(A, B)** Projected degradation of estimation precision (*σ*_AR_ and *σ*_MR_) as a continuous function of uniformly applied baseline measurement noise scalars, plotted across specific observational temporal densities (*N ∈ {*10, 25, 50 *}*). **(C)** Non-linear decay curves outlining estimation uncertainty strictly against the sequential measurement sample dimension (*N≤* 100) evaluated at the exact nominal baseline theoretical noise profile (1.0 *×*). The horizontal dashed boundary denotes the optimal clinical target boundary of 5% estimation uncertainty.

Even under elevated measurement noise (4.0 *×* multiplier), *N* = 50 physiologically varied samples remained sufficient to hold both fractions inside the 5% clinical margin (*σ*_AR_ = 0.035; *σ*_MR_ = 0.047). Across all noise levels, MR required marginally more observations to reach the same precision as AR, consistent with its attenuated univariate observability (Figure 1).

### 3.3 Feature Ablation and Theoretical Information Profiling

To identify which features carry the valvular information, we ablated each feature in turn by inflating its noise variance tenfold (Section 2.7). Baseline performance on *N* = 50 physiological samples was *σ*_AR_ = 0.0088 and *σ*_MR_ = 0.0119.

The profiles revealed that inversion accuracy depends on a small subset of reflection- and ventriculoarterial-coupling markers (Figure 3). Dicrotic notch timing was informative for both lesions: ablating it increased the bounds to *σ*_AR_ = 0.0169 (+0.0081) and *σ*_MR_ = 0.0184 (+0.0065). Augmentation index was uniquely critical for MR, with its ablation nearly doubling the theoretical bound to *σ*_MR_ = 0.0218 (+0.0100) while leaving AR estimation essentially untouched.

**Figure 3:**
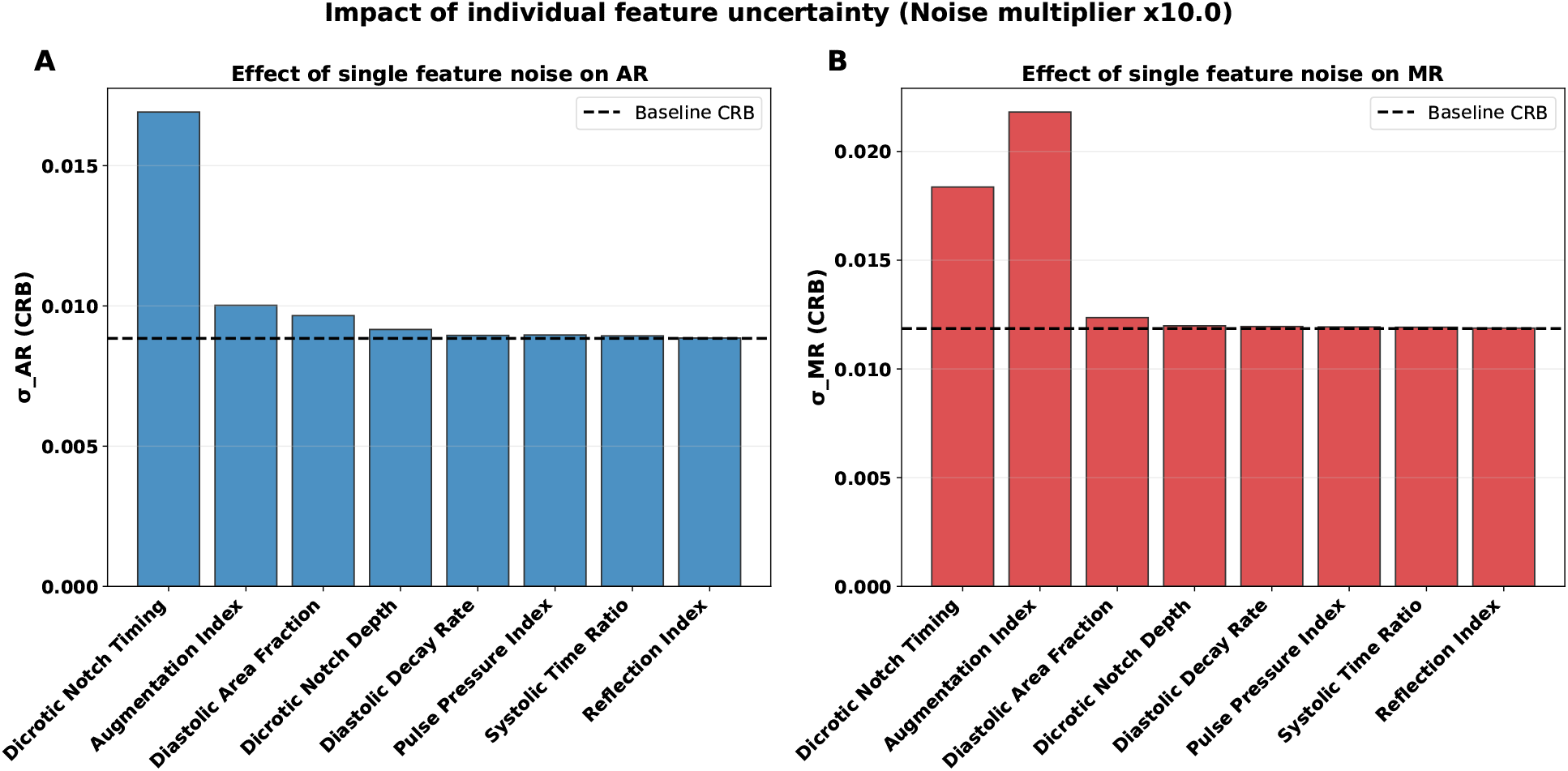
Feature ablation profiling characterizing theoretical observability limits via the Cramér-Rao Bound (CRB). Relative dependence on PPG features is quantified by measuring the deterioration of overall estimation precision of AR and MR fraction when individual peripheral waveform features are systematically penalized with high-noise. Each colored continuous bar represents the theoretical standard deviation (*σ*) limit for inferring **(A)** aortic regurgitation (AR) and **(B)** mitral regurgitation (MR) when subjecting that specific feature to very high measurement noise (10.0 *×* multiplication of characteristic baseline variance). Evaluations are mapped against a uniformly varied sampling background (*N* = 50 distinct background physiological states). The horizontal dashed black lines indicate the bounded precision inherent to the normative noise conditions for all PPG features. Notably, bounds are less tolerant to noise in Dicrotic Notch Timing across both pathologies, while identifying Mitral Regurgitation as also relying on Augmentation Index.

Geometric diastolic markers (diastolic area fraction and dicrotic notch depth) provided modest but non-negligible information (+0.0008 and +0.0003 for AR). Ablation of the remaining features (diastolic decay rate, pulse pressure index, systolic time ratio, reflection index) produced negligible degradation (*≤* +0.0001). These results indicate that a small subset of features carries most of the valvular information, although the full set remains useful for constraining the nuisance parameters *θ*_circ_ in the partially-observed joint inverse problem.

### 3.4 Surrogate Neural Network Performance

We trained a neural surrogate of the 1D hemodynamic model to map a 14-dimensional parameter vector—four valvular regulators (aortic and mitral regurgitant fractions, the AR diastolic decay constant *τ*, and the MR systolic onset delay) together with ten systemic circulation parameters—to the eleven PPG features (Section 2.8). Held-out performance on 5,000 test samples is reported in Table 2. All features achieved *R*^2^ *>* 0.84, with most exceeding 0.9; mean absolute percentage errors remained below 12% for features whose values are bounded away from zero, and predicted means agreed closely with ground truth, indicating negligible bias. We use the surrogate only in the downstream Bayesian inference loop to evaluate posteriors efficiently (Sections 2.10 and 2.11); the ground-truth synthetic measurements used during longitudinal tracking are always generated from the full 1D model.

**Table 2:** Neural network surrogate performance on test set with 5000 samples. Note that MAPE is not reported for Augmentation index and Dicrotic notch depth since they consist of values near zero or cross zero which causes extreme MAPE (%) measures with small values.

| Feature | $R^2$ | MAPE (%) | Mean (True) | Mean (Pred) |
| --- | --- | --- | --- | --- |
| Pulse transit time | 0.9226 | 1.88 | 64.8954 | 64.7452 |
| Pulse wave velocity | 0.8744 | 3.77 | 15.3453 | 15.5027 |
| Augmentation index | 0.8987 | — | -0.1878 | -0.1818 |
| Reflection index | 0.9178 | 1.19 | 0.6731 | 0.6748 |
| Systolic time ratio | 0.9551 | 2.45 | 0.3452 | 0.3436 |
| Dicrotic notch timing | 0.8477 | 8.32 | 200.9055 | 200.1539 |
| Diastolic decay rate | 0.9654 | 11.71 | 4.2774 | 4.2634 |
| Dicrotic notch depth | 0.8779 | — | 0.4922 | 0.4856 |
| Diastolic area fraction | 0.9513 | 3.06 | 0.8164 | 0.8201 |
| Pulse pressure index | 0.9670 | 1.07 | 1.8990 | 1.8952 |

### 3.5 Longitudinal State Tracking Under Dynamic Clinical Scenarios

We next evaluated the full MCMC-Kalman framework (Section 2.9) on a curated set of synthetic trajectories that reproduce canonical valvular disease dynamics (Section 2.10). Performance was quantified by the root-mean-square error (RMSE) and the maximum absolute deviation (MaxErr) between the filter’s posterior mean and the ground-truth regurgitant fraction.

Under steady, constant-velocity progression (Figure 4A-D), the filter recovered the underlying regurgitant fractions even when all eight systemic parameters were constrained only by their population-level physiological bounds (Bounds in Table 1). Aortic regurgitation tracked more accurately than mitral regurgitation across all conditions. The Stable AR scenario attained RMSE = 0.0381 (MaxErr = 0.0582), improving on Stable Primary MR (RMSE = 0.0441, MaxErr = 0.0778); the High-Risk AR profile (*v*_0_ = 0.004/month) achieved the lowest error overall (RMSE = 0.0274); and Secondary MR showed the widest excursion (MaxErr = 0.0834).

**Figure 4:**
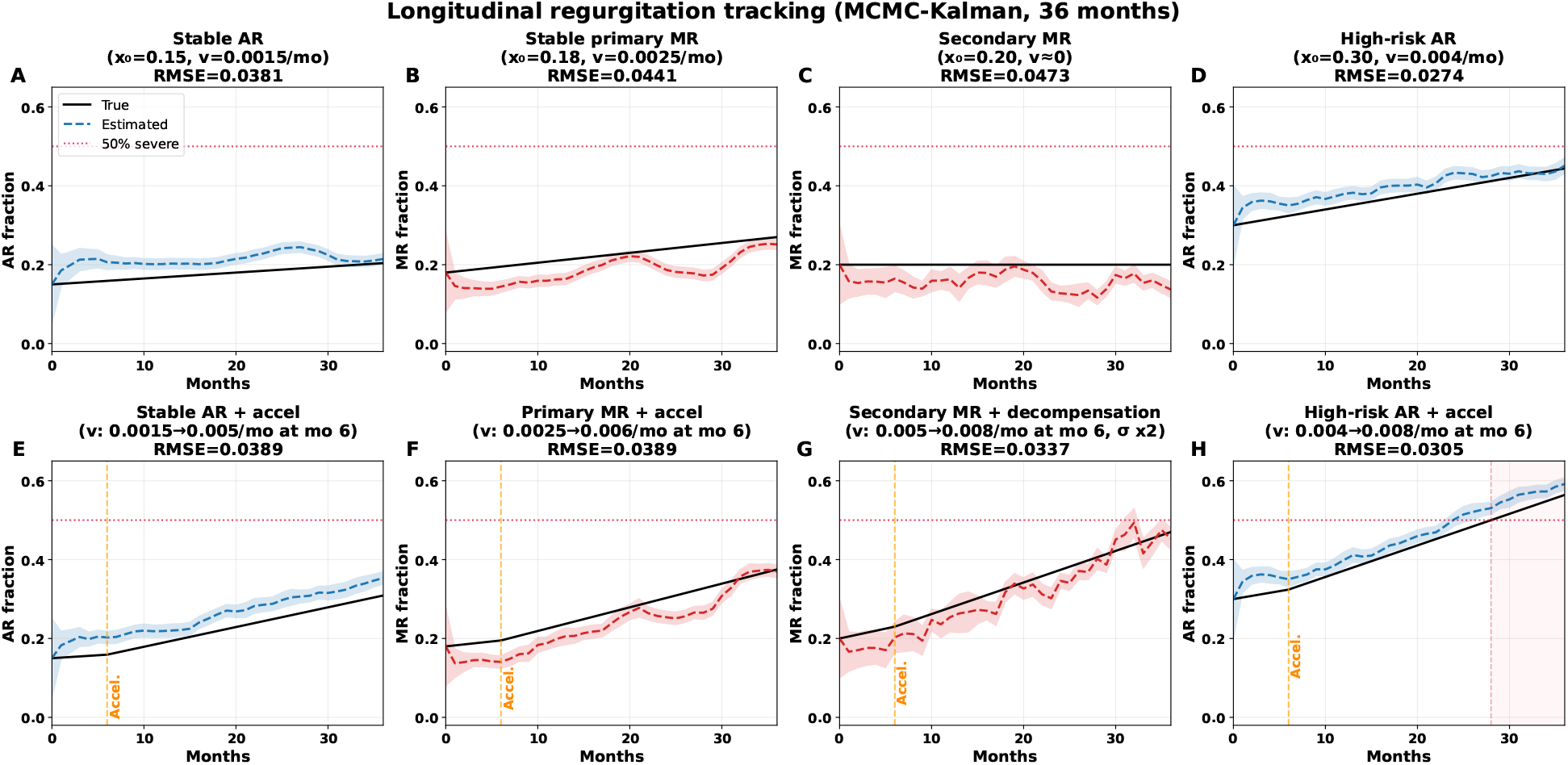
Longitudinal tracking of Aortic (AR) and Mitral (MR) regurgitation severity over a 36-month horizon using the MCMC-Kalman framework. The true underlying regurgitant fraction (solid black line) is estimated sequentially from peripheral PPG features. Filter state estimates are presented as dashed lines (blue for AR tracking, red for MR tracking) bounded by shaded credible intervals. A clinically relevant severe regurgitation threshold of 50% (0.5 fraction) is denoted by the horizontal dotted red line. The root mean square error (RMSE) for each trajectory is provided in the panel titles. **(A–D) Constant Progression Dynamics:** The top row evaluates filter lock-on and tracking under continuous baseline progression. **(A)** Chronic stable AR characterized by low baseline and slow clinical progression (*x*_0_ = 0.15, *v* = 0.0015*/*month). **(B)** Stable primary MR (*x*_0_ = 0.18, *v* = 0.0025*/*month). **(C)** Load-dependent secondary MR lacking active structural progression (*x*_0_ = 0.20, *v ≈* 0). **(D)** High-risk AR undergoing faster remodeling dynamics (*x*_0_ = 0.30, *v* = 0.004*/*month). **(E–H) Accelerated Progression and Decompensation:** The bottom row evaluates the robustness and responsiveness of the filter against dynamic shifts in progression (indicated by the vertical dashed yellow line marked “Accel.” at month 6). **(E)** Stable AR accelerating abruptly (drift *v* escalating from 0.0015 to 0.005*/*month). **(F)** Primary MR subjected to an acceleration event mimicking a rapid deterioration such as chordal rupture or active remodeling (*v*: 0.0025 *→* 0.006*/*month). **(G)** Secondary MR undergoing clinical decompensation, formulated as an onset of significant worsening (*v*: 0.005 *→* 0.008*/*month) coupled inherently with a doubling of the population progression uncertainty covariance (*σ ×* 2). **(H)** High-risk AR experiencing severe, escalated remodeling (*v*: 0.004 *→* 0.008*/*month), smoothly tracking as the disease severity crosses into the pathological *≥* 50% criteria.

To probe responsiveness to abrupt clinical events, we injected sudden acceleration in disease velocity at month six (Figure 4E-H), mimicking flail leaflets or acute chordal rupture. The EMA-based growth-rate estimator adapted to the new slopes without runaway divergence: Stable AR and Primary MR with delayed failure were both tracked to RMSE = 0.0389, and simulated massive Secondary MR decompensation remained bounded (RMSE = 0.0337, MaxErr = 0.0642).

### 3.6 Population-scale tracking validation

To assess generalization across clinically heterogeneous patients, we scaled the analysis to eight pre-specified progression scenarios, each enrolling *N* =30 synthetic patients followed monthly over a 36-month horizon (Section 2.11). Population results are summarized in Table 3 and Figure 5. AR progression was tracked with sub-0.03 mean RMSE under both stable and high-risk priors, with or without a progressive hypertension confounder. MR tracking was feasible but exhibited larger variance and a systematic under-estimation bias at low-to-moderate regurgitant fractions (Figure 5).

**Table 3:** Population-scale MCMC-Kalman regurgitation tracking accuracy across eight clinical scenarios (*N* =30 patients per scenario, 36-month horizon). Mean RMSE *±* standard deviation reported in regurgitant fraction units. Corresponds to aggregate measures from Figure 5.

| Scenario | Valve | Confounder | RMSE (mean ± std) |
| --- | --- | --- | --- |
| Stable AR | AR | — | $0.0284 \pm 0.0135$ |
| Stable AR + HTN | AR | HTN | $0.0265 \pm 0.0135$ |
| High-risk AR | AR | — | $0.0229 \pm 0.0106$ |
| High-risk AR + HTN | AR | HTN | $0.0223 \pm 0.0116$ |
| Stable MR | MR | — | $0.1009 \pm 0.0719$ |
| Stable MR + HTN | MR | HTN | $0.0826 \pm 0.0629$ |
| High-risk MR | MR | — | $0.0891 \pm 0.0570$ |
| High-risk MR + HTN | MR | HTN | $0.0752 \pm 0.0410$ |

**Figure 5:**
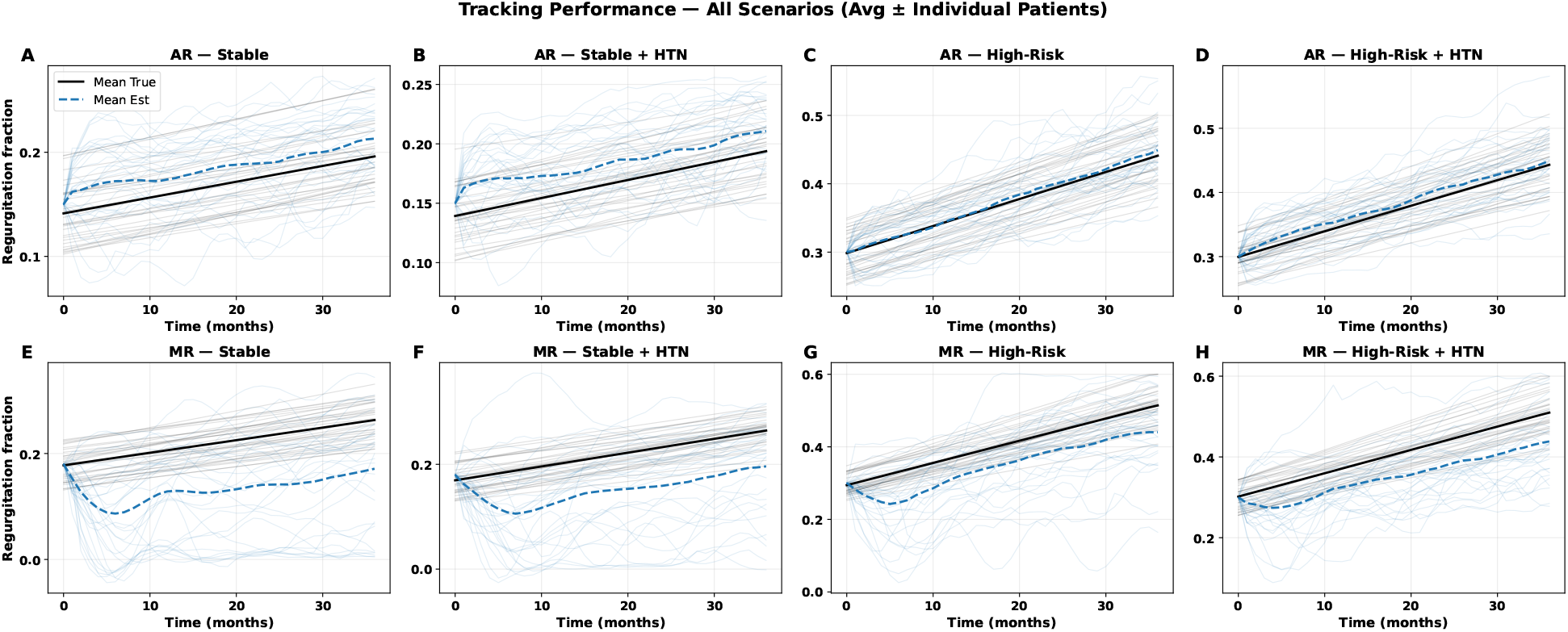
Population-scale tracking performance across all eight clinical scenarios (*N* =30 patients per scenario, 36-month horizon). Each panel displays individual patient ground-truth trajectories (gray) and filter estimates (light blue), overlaid with the population-mean true path (solid black) and mean estimate (dashed blue). Patient-specific initial severities *x*_0_ and progression rates *v*_0_ were sampled from scenario-level prior distributions, while the Kalman filter was initialized at the unperturbed population-average prior to stress-test convergence. **Top row (AR):** Aortic regurgitation tracking under stable (*x*_0_=0.15, *v*_0_=0.0015/month) and high-risk (*x*_0_=0.30, *v*_0_=0.004/month) progression regimes, with and without a progressive hypertension confounder (∆MAP=+10–30 mmHg, ∆*C*_wk_= *−* 10–20% over 36 months). The mean estimated trajectory converges tightly to the true path in all four AR panels within the first 3–5 months, with residual bias remaining below 0.03 regurgitant fraction units. **Bottom row (MR):** Mitral regurgitation tracking under the same scenario structure. A systematic negative bias in the mean estimate is evident across all MR panels, particularly during the first 10–15 months, reflecting the lower sensitivity of peripheral PPG features to mitral valve pathology. Progressive hypertension partially mitigates this bias by introducing additional hemodynamic variation that improves posterior resolution of *f*_MR_. Scenario-level RMSE (mean *±* std) is reported in Table 3.

#### 3.6.1 Aortic Regurgitation (AR)

AR fraction was tracked with high fidelity in all four AR-containing scenarios. Stable AR (*x*_0_=0.15, *v*_0_=0.0015/month, *σ*_path_=0.006/month) yielded RMSE = 0.0284 *±* 0.0135, with population-mean estimates converging to the ground-truth trajectory within the first three to five months (Figure 5A). High-risk AR (*x*_0_=0.30, *v*_0_=0.004/month, *σ*_path_=0.010/month) produced the best AR accuracy (RMSE = 0.0229 *±* 0.0106), likely because larger excursions from baseline amplify the morphological PPG changes that carry information about the regurgitant fraction (Figure 5C). Adding a progressive hypertension confounder had negligible impact: stable AR+HTN gave RMSE = 0.0265 *±* 0.0135 (Figure 5B) and high-risk AR+HTN gave RMSE = 0.0223 *±* 0.0116 (Figure 5D). This insensitivity indicates that the joint posterior over *θ*_circ_ absorbs slow drifts in peripheral compliance and mean arterial pressure without contaminating the AR estimate. Across all four AR scenarios the mean tracking error remained below 0.03 regurgitant fraction units.

#### 3.6.2 Mitral Regurgitation

MR tracking was systematically less accurate than AR tracking. Stable MR (*x*_0_=0.18, *v*_0_=0.0025/month) yielded RMSE = 0.1009 *±* 0.0719 and displayed a persistent negative bias: the population-mean estimate consistently underestimated the true trajectory during the first ten months before partially recovering (Figure 5E). High-risk MR (*x*_0_=0.30, *v*_0_=0.006/month, *σ*_path_=0.010/month) improved modestly to RMSE = 0.0891 *±* 0.0570, consistent with the larger signal-to-noise ratio at elevated regurgitant fractions (Figure 5G).

Unexpectedly, the progressive hypertension confounder improved—rather than degraded—MR tracking relative to the corresponding normotensive scenarios. Stable MR+HTN reduced RMSE from 0.1009 to 0.0826 *±* 0.0629 (*−*18%; Figure 5F) and high-risk MR+HTN reduced RMSE from 0.0891 to 0.0752 *±* 0.0410 (*−*16%; Figure 5H, Table 3). We interpret this as evidence that gradually rising mean arterial pressure and falling peripheral compliance enlarge the hemodynamic excitation range, which in turn sharpens the MR posterior by driving the operating point away from the low-contrast region where peripheral PPG features are least sensitive to mitral pathology.

### 3.7 Systemic Parameter Identifiability and Its Relation to Tracking Accuracy

Because the filter must infer eight systemic parameters (*L, R*_1_, *R*_2_, *C*_wk_, *r*_0,*p*_, *r*_0,*d*_, *β*_*p*_, *β*_*d*_) jointly with the regurgitant fraction, we asked how posterior identifiability of these nuisance parameters relates to downstream tracking accuracy. Figure 6 plots the time-evolution of each parameter’s relative posterior standard deviation (normalized to its value at *t* = 0, left axis) alongside the Kalman tracking uncertainty *σ* on the regurgitant fraction (right axis) for the two worst-performing scenarios overall, stable AR and stable MR. Within each row, the left panel shows the patient tracked most accurately (lowest RMSE; A, C) and the right panel the patient tracked least accurately (highest RMSE; B, D). Values below one indicate posterior contraction relative to the prior; sustained values near or above one indicate that the data did not resolve that parameter.

**Figure 6:**
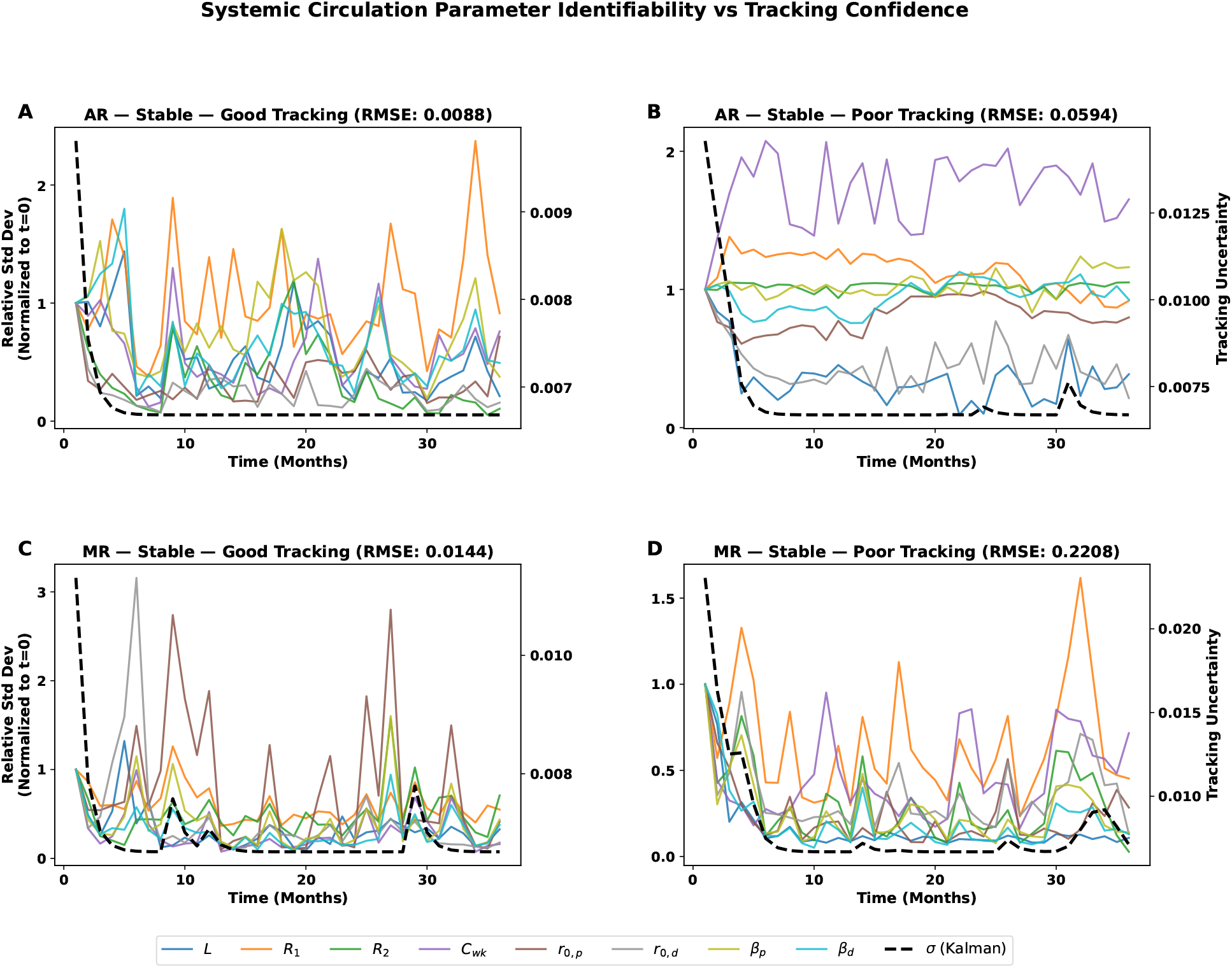
Systemic circulation parameter identifiability versus Kalman tracking confidence for stable aortic regurgitation (AR) and mitral regurgitation (MR). Each panel shows the evolution of the relative posterior standard deviation (left axis, normalized to *t* = 0) for eight systemic circulation parameters alongside the Kalman-filter tracking uncertainty *σ* (right axis, dashed). The top row corresponds to the stable AR scenario and the bottom row to the stable MR scenario; within each row the left panel depicts the best-tracked patient (lowest RMSE) and the right panel the worst-tracked patient (highest RMSE). A rapid collapse of *σ* in early months reflects Kalman convergence, while systemic circulation parameter standard deviations that remain elevated or oscillatory indicate limited posterior identifiability for those parameters under that tracking regime.

For both AR and MR, the Kalman uncertainty *σ* collapses within two to five months, but the systemic parameters do not contract monotonically. Some parameters tighten quickly, while others oscillate or even widen as the filter encounters new measurements, consistent with shallow likelihood ridges in which different *θ*_circ_ combinations generate nearly identical PPG features.

To quantify this relationship across the full cohort, we regressed patient-level RMSE on the time-averaged relative posterior standard deviation of each systemic parameter (Figure 7). All eight parameters exhibited negative correlations with RMSE; peripheral compliance *C*_wk_ (panel D) and proximal baseline radius *r*_0,*p*_ (panel E) reached statistical significance (Pearson *r* = *−*0.09, *p* = 0.003 and *r* = *−*0.21, *p* = 0.001, respectively). In other words, patients whose systemic parameters were *more* tightly constrained during inference tended to have *worse* regurgitant-fraction tracking. This apparent paradox is consistent with the shallow-ridge picture above: when the sampler collapses onto a narrow but incorrect region of *θ*_circ_ space, the regurgitant-fraction estimate inherits a systematic bias, whereas appropriately diffuse posteriors over the nuisance parameters leave room for the Kalman filter to correct the valvular estimate as new data arrive.

**Figure 7:**
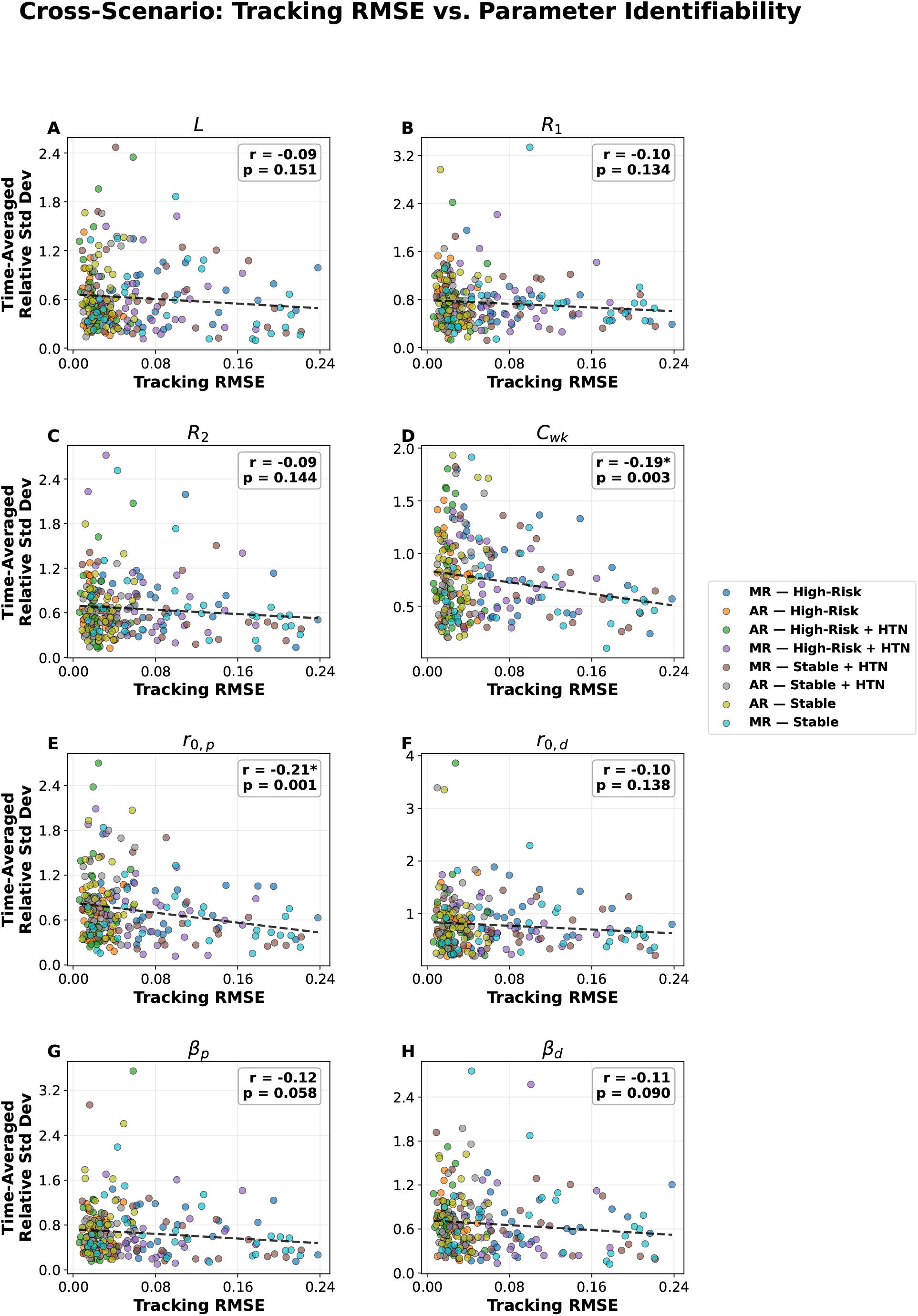
Cross-scenario correlation between bio-parameter identifiability and tracking performance. **(A-H)** Scatter plots demonstrating the relationship between the time-averaged relative standard deviation (a proxy for uncertainty) of eight key systemic circulation bioparameters and the patient-specific longitudinal tracking error (RMSE) computed over a 36-month horizon. Each data point represents an individual patient’s trajectory. Marker color indicates

## 4 Discussion

This study establishes, in silico, that the longitudinal progression of aortic and mitral regurgitation fractions can be recovered from peripheral photoplethysmography morphology despite unknown and drifting background hemodynamics. Three findings anchor this conclusion. First, the morphology of the distal PPG waveform carries a quantifiable, valve-specific signature of regurgitation severity, strongly so for AR and more subtly for MR. Second, under realistic intra-individual physiological variability, the Cramér–Rao bound indicates that as few as *N ≈* 50 monthly measurements suffice to drive the theoretical tracking uncertainty below 1.2% regurgitant-fraction units for both lesions. Third, the MCMC–Kalman framework approaches this theoretical bound in practice for AR (RMSE *≤* 0.028 across four scenarios, including a progressive hypertension confounder) and reaches moderate fidelity for MR (RMSE 0.075–0.101), while simultaneously marginalizing over eight unknown systemic circulation parameters.

The asymmetry between AR and MR observability is a direct consequence of where each lesion perturbs the aortic inlet. Aortic regurgitation operates *within* the systemic circulation: the incompetent aortic valve couples diastolic aortic pressure back to the left ventricle, producing both a pathologically augmented forward ejection and an exponential diastolic runoff that widens pulse pressure, blunts the dicrotic notch, and stiffens the effective proximal impedance [5, 6]. Each of these modifications leaves a mechanical fingerprint that propagates distally through the 1D arterial tree, which is why pulse-wave velocity, stiffness index, augmentation index, and dicrotic notch timing all carried SNRs above 10 for AR (Figure 1). Mitral regurgitation, by contrast, acts *upstream* of the aortic valve: ventricular volume lost to the left atrium is compensated in part by faster, earlier systolic ejection, so the forward aortic waveform is reduced in amplitude and shortened in duration but otherwise retains a normal shape. The peripheral footprint of MR is therefore smaller and more collinear with ordinary stroke-volume variation. This physiological intuition aligns with classical hemodynamic theory [34] and is consistent with the narrower set of MR-informative features identified by our ablation study — essentially augmentation index and, to a lesser extent, dicrotic notch timing and pulse pressure index (Figure 3).

Physiological variability, rather than being a nuisance, is a structural requirement for identifiability. The Cramér– Rao analysis shows that a single-shot measurement cannot disentangle the regurgitant fraction from the eight latent circulatory parameters *θ*_circ_: the forward map is locally degenerate. However, as heart rate, mean arterial pressure, and vascular properties fluctuate naturally from day to day, each measurement samples a slightly different slice of the forward map, and the aggregate Fisher information accumulates rank along axes that are otherwise invisible. This is precisely the rationale underpinning sequential Bayesian and variational data-assimilation approaches in computational hemodynamics [28, 29, 31, 30, 32], and it explains why the addition of a progressive hypertension confounder — which increases the hemodynamic excitation observed by the tracker — paradoxically *improved* MR tracking RMSE by 16–18% rather than degrading it (Table 3). The same mechanism suggests that richer ecological data from wearables, which naturally sample a wide physiological envelope across activities of daily living, may be more informative than the narrow, rest-only snapshots traditionally collected in the clinic.

The identifiability–performance relationship (Figure 7) surfaces a subtle failure mode of joint Bayesian inversion that deserves explicit emphasis. Counter to the intuition that tightening posterior uncertainty is always beneficial, we observed significant *negative* correlations between the time-averaged posterior standard deviation of latent parameters (notably peripheral compliance *C*_wk_ and proximal radius *r*_0,*p*_) and tracking RMSE: patients whose latent parameters were most tightly pinned down also suffered the largest tracking errors. This behavior is consistent with a well-known pathology of structurally non-identifiable models: when the likelihood surface has a shallow ridge, MCMC walkers can migrate into a narrow but incorrect neighborhood of parameter space and then become stuck there, propagating a confidently wrong belief about *θ*_circ_ into the regurgitation estimate [33, 32]. Healthy posteriors in this setting should remain partially diffuse along the non-identifiable directions; over-contraction is a symptom, not a success. Practical mitigations include multi-start re-seeding of the walker ensemble, informative hyperpriors from wearable-derived surrogates for heart rate and blood pressure [12, 13, 14], and the inclusion of additional morphological features that break the residual degeneracies.

Situated in the broader landscape of digital cardiology, this work contributes a physics-based scaffold for a *valvular* digital twin [35] powered by the signal most densely available in consumer and clinical settings [8, 10, 37]. Complementary AI-ECG approaches have shown that surface electrocardiograms carry discriminative information about moderate-to-severe left-sided valvular disease [20]; our results indicate that the peripheral pulse wave carries graded, quantitative information about the *severity* of the same lesions, enabling tracking rather than mere classification. Because PPG and ECG are routinely co-acquired in modern wearables, the two modalities are natural candidates for fusion, and the Bayesian framework presented here admits additional likelihood terms with no structural change. Looking forward, such a fused, continuously updated estimate of regurgitant fraction could complement the episodic echocardiographic surveillance recommended by current guidelines [7, 5, 6], flagging accelerations of disease between clinic visits and informing the timing of definitive intervention.

Several limitations of the present work bound its interpretation. First, all results are synthetic: both the training data for the surrogate and the “observations” processed by the tracker were produced by the same 1D solver. Model-to-model experiments establish an upper bound on achievable performance, but cannot substitute for prospective validation against paired echocardiographic and wearable recordings in human cohorts. Second, the arterial domain is a single-path tube from the aortic root to the pedal digital artery. While such reduced models reproduce the dominant features of peripheral waveforms and have been repeatedly validated [22, 26, 27], a branched anatomy with multiple terminal beds [25] may change the quantitative sensitivity of specific features and open additional, site-specific diagnostic axes (for example, comparing finger-to-ear or finger-to-toe transit). Third, we treated the optical PPG signal as a noisy linear observation of the distal cross-sectional area; real wearable signals are further corrupted by motion artifact, contact pressure, skin tone, and perfusion variation, and the signal-to-noise ratios we simulated should therefore be regarded as optimistic [10]. Fourth, valve regurgitation was represented purely as a parametric modification of the aortic inlet waveform rather than through a coupled left-heart model; chronic remodeling of the ventricle, atrium, and circulatory impedance was approximated through scalar compensation laws. A fully coupled ventricular–arterial formulation would permit explicit representation of eccentric hypertrophy, afterload-dependent ejection, and rhythm interactions that are known to modulate peripheral waveform morphology. Finally, we simulated pure-AR and pure-MR trajectories; mixed valvular disease and concurrent stenotic lesions, which are common in aging populations [2, 3], were not explored and would likely require expansion of the inference dimensionality and the feature set.

Future work will pursue three directions. First, prospective collection of simultaneous high-fidelity PPG and echocardiographic measurements in patients with graded AR and MR severity, across multiple follow-up visits, is needed to convert the present upper bound into clinically calibrated performance. Regulatory-grade in-silico-to-in-vivo credibility assessment of the surrogate and tracker along the lines proposed by the ASME V&V 40 framework [38] would support eventual deployment. Second, the forward model can be enriched by replacing the single compliant tube with a branched arterial network [25] and by coupling a reduced-order ventricular–valvular model to capture the hemodynamic consequences of remodeling explicitly. Third, the inference engine can be expanded to absorb complementary signals routinely available in wearables — single-lead ECG, ballistocardiography, seismocardiography, and cuff-less blood pressure estimates — through additional likelihood terms, and to support active learning strategies that request a clinic-grade echocardiogram only when predicted uncertainty exceeds a decision-relevant threshold.

## 5 Conclusion

We have shown, through a fully synthetic but physiologically grounded pipeline, that peripheral photoplethysmography carries sufficient information to track the longitudinal progression of aortic and mitral regurgitation severity, provided the inverse problem is framed as a joint Bayesian inference over valvular and systemic circulation parameters. A 1D Navier–Stokes arterial model with parametric AR and MR inlet waveforms, combined with a trained neural surrogate and an MCMC–Kalman tracking framework, delivered RMSE of 0.023–0.028 regurgitant-fraction units for AR and 0.075–0.101 for MR across eight canonical clinical scenarios, including progressive hypertension confounders, over a 36-month horizon. Theoretical analyses via the Cramér–Rao bound showed that dicrotic notch timing and augmentation index are the dominant informative features, and that natural day-to-day physiological variability is an asset rather than a confounder, anchoring the identifiability of latent circulatory parameters through repeated measurement. Taken together, these results provide a computational foundation for wearable, physics-informed surveillance of valvular heart disease and motivate prospective clinical studies designed to calibrate and validate the framework against echocardiographic ground truth.

## Data Availability

All code & data produced in the present study are available upon reasonable request to the authors.

